# Deep Learning Prediction of Left Atrial Structure and Function from 12-lead Electrocardiograms

**DOI:** 10.1101/2025.09.29.25336490

**Authors:** Jennifer A Brody, Vidhushei Yogeswaran, Kerri L Wiggins, Colleen M Sitlani, Joshua C Bis, Lin Yee Chen, Susan R Heckbert, João A C Lima, W T Longstreth, Elsayed Z Soliman, Geoffrey H Tison, Ting Ye, Bruce M Psaty, Ali Shojaie, James S Floyd

**Affiliations:** Cardiovascular Health Research Unit, Department of Medicine, University of Washington, Seattle, WA, USA; Department of Epidemiology, University of Washington, Seattle, Washington, USA; Lillehei Heart Institute & Cardiovascular Division, Department of Medicine, University of Minnesota Medical School, Minneapolis, MN, USA; Division of Cardiology, Department of Medicine, Johns Hopkins University School of Medicine, Baltimore, MD, USA; Department of Neurology, University of Washington, Seattle, Washington, USA; Department of Cardiovascular Medicine, Wake Forest University School of Medicine, Winston-Salem, North Carolina, USA; Division of Cardiology, University of California-San Francisco, San Francisco, California, USA; Department of Biostatistics, University of Washington, Seattle, WA, USA; Department of Health Systems and Population Health, University of Washington, Seattle, WA, USA; Department of Statistics, University of Washington, Seattle, WA, USA

## Abstract

Abnormal cardiac atrial structure and function (atrial cardiopathy)^1^ typically precedes atrial fibrillation (AF) and predicts other cardiovascular complications, yet detection is limited by the cost and limited accessibility of high-quality cardiac imaging. We trained a deep learning model (ECG-AI) using 12-lead electrocardiograms paired with 21,749 cardiac magnetic resonance scans to predict left atrial structure and function. ECG-AI measures of atrial cardiopathy were strongly associated with new-onset AF, heart failure, and ischemic stroke after adjustment for clinical risk factors and biomarkers in two external cohorts, outperforming imaging measures and clinical risk factors. The risk of cardioembolic stroke, the hallmark complication of AF, was 66% greater per SD left atrial volume. In a screening population, ECG-AI predicted subclinical AF from 14-day cardiac monitoring better than a clinical risk prediction tool. Our ECG-AI is an inexpensive, accessible tool that identifies individuals at high-risk for AF and related complications.

## Introduction

Growing evidence suggests that abnormalities in the structure and function of the cardiac atrial are independent risk factors for AF, heart failure, and stroke, even among those without a clinical diagnosis of AF^2–4^. Most studies on atrial cardiopathy have focused on widely available electrocardiographic or echocardiographic or measures of left atrial size, which have poor sensitivity^1,5–7^. Cardiac magnetic resonance imaging (CMR) is the gold-standard imaging modality for assessing atrial volume and function^8^, but is not suitable as a screening tool because of high costs and limited availability^9,10^.

Recent studies have applied artificial intelligence (AI) and deep learning models to the resting 12-lead electrocardiogram (ECG), which has moderate accuracy in predicting short-and long-term risks of AF.^11–13^ The clinical diagnosis of AF is influenced by the presence of symptoms and the duration of episodes, as well as characteristics that have little relation to biological risk such as race^14,15^. This ascertainment bias fundamentally constrains the ability of models trained on clinically diagnosed AF to identify the underlying atrial substrate that predisposes to AF. The development and validation of models that predict left atrial and structure and function from ECG data has the potential to translate atrial cardiopathy into a tractable target for cardiovascular disease prevention, including application for population-wide screening for AF.

In this manuscript, we present a deep learning approach that reliably predicts left atrial volumes and ejection fraction from the resting 12-lead ECG. Our ECG-AI models, trained on data with high-quality CMR imaging from over 20,000 participants and externally validated in two large population-based cohort studies, correlates well with direct imaging measures and are independently associated with new-onset AF, stroke, and heart failure. More importantly, this ECG-AI outperforms currently available clinical risk models and existing imaging measures of atrial structure and function and AF. We also conducted genetic correlation analyses and model explainability studies to characterize the biological basis and ECG features underlying our model’s predictions. This approach offers a translatable, low-cost method for assessing atrial size and function using the widely available ECG, facilitating the widespread screening of individuals who may be at high-risk for AF-related consequences across diverse clinical settings.

## Results

### ECG-AI model

We developed a deep learning model to predict left atrial imaging measures derived from CMR using digital 10-second 12-lead ECGs performed on the same day as the CMR imaging studies in the UK Biobank (UKB) participants. Left atrial minimal and maximal volumes (LAVmin, LAVmax) and total left atrial ejection fraction (LAEF) were extracted from CMR images using a validated machine-learning-based analysis pipeline^16^. Unique participants with concurrent ECGs and CMR scans that passed QC (N=48,300) were randomly split into 45% training, 5% validation, and 50% test samples (**Supplemental Figure 1**). Participants in the training sample were 53% female, with a median age of 65 years, and a low prevalence of heart failure (0.4%) and prior myocardial infarction (2.1%) (**Supplemental Table 1 and 2**). Participants in the test sample had similar characteristics. In the training sample, we used the median beat of the eight non-derived leads of the 12-lead ECGs to train several models with a range of hyperparameters to predict LAVmin, LAVmax and LAEF (**Figure 1**). The validation sample was used for model selection and hyperparameter tuning. For each measure, predictions from the five highest-performing models were ensembled to create the final ECG-AI prediction model. Additional information on model architecture and training is in the Online Methods and final parameters are provided in **Supplemental Data Table 3**. The resulting ECG-AI model was then evaluated in the testing sample. The predicted (ECG-AI) LA measures were correlated with the imaging (CMR) measures in the testing partition: r = 0.50 for LAVmin and LAVmax and 0.40 for LAEF (**Figure 1)**.

**Figure 1:**
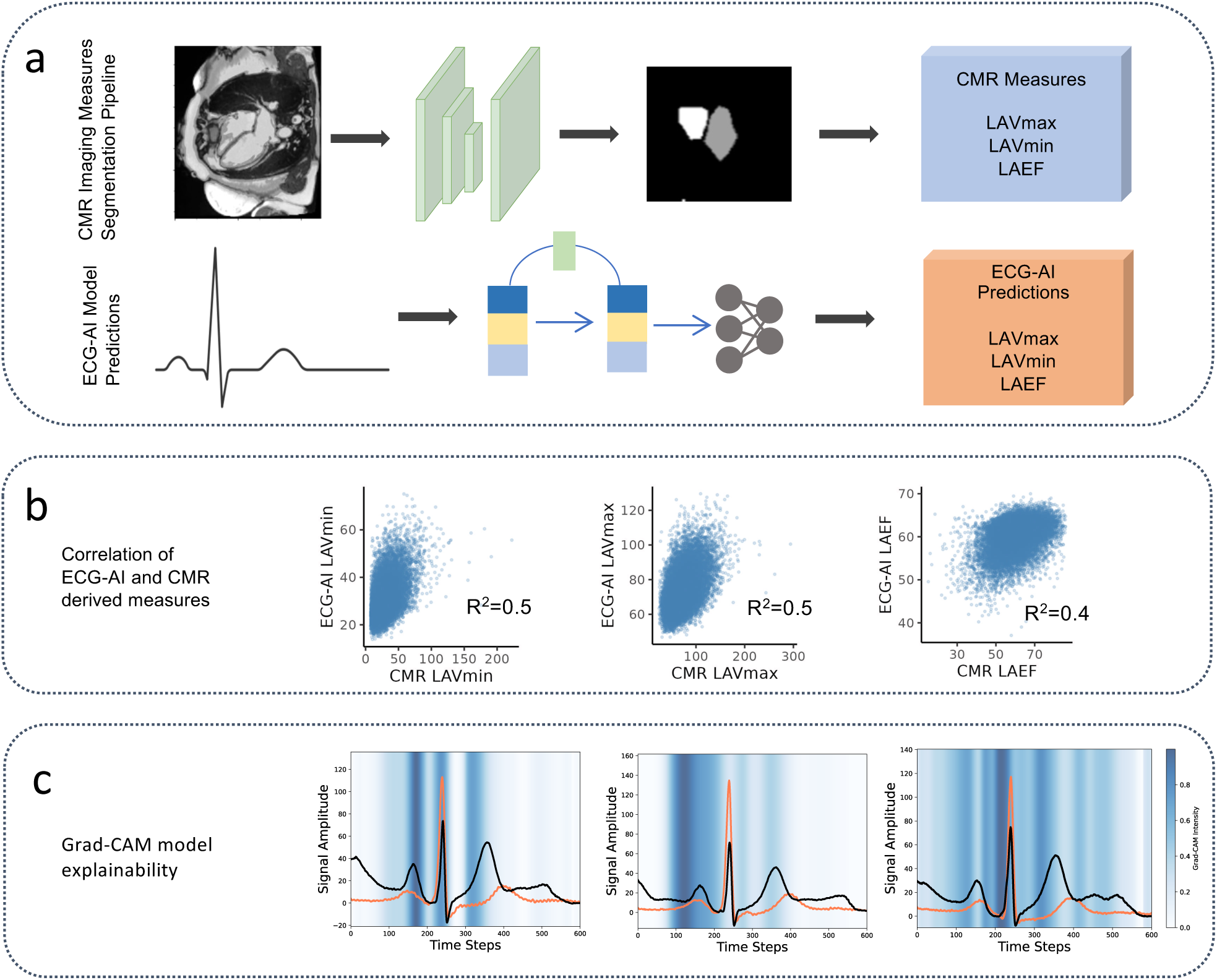
ECG-AI model development and evaluation. a) Atrial volumes were derived from CMR images in UKB by applying a deep-learning model to segment the left atrial minimum and maximum volumes. We then trained our ECG-AI models to predict these volumes (LAVmin, LAVmax) and their proportion (LAEF) from 12-lead ECGs at the same study visit. b) In the test partition of UKB, the Pearson correlations between the segmented CMR LA measure and ECG-AI predicted LA measure were 0.50 for LAVmin and LAVmax and 0.40 for LAEF. c) Heatmaps for the ECG regions driving the predictions of the ECG-AI in UKB. The blue intensity corresponds to feature importance, with the darkest blue signaling the highest importance. The 50 largest volumes (worse) for LAVmin and the 50 smallest ejection fractions for LAEF (worse) median beats are averaged and displayed in orange. For comparison, the average of the 50 smallest (volumes) or largest (LAEF) are plotted in black. For LAVmin, the model is primarily focused on the decline of the P-wave, while LAVmax is focused on the first half of the P-wave. In contrast, the LAEF signal is focused on the PQ segment. Images reproduced by kind permission of UK Biobank ©

### ECG-AI associations with clinical outcomes

In the UKB test sample, ECG-AI and CMR atrial measures were evaluated for associations with clinical outcomes. There were 331 AF, 71 ischemic stroke, and 118 heart failure events during a mean follow-up of 4 years. Left atrial volumes were indexed to body surface area (BSA) to obtain LAVImin and LAVImax. After adjustment for established clinical risk factors, CMR-derived LA measures were significantly associated with all outcomes (**Table 1**). Incident AF hazard ratios (HRs) for ECG-AI measures were slightly attenuated: each standard deviation (SD) higher (worse) CMR LAVImin was associated with a 44% increased risk (95% CI 34-53%) while ECG-AI-predicted LAVImin was associated with a 37% increased risk (95% CI 26-50%). In contrast, PR interval from the 12-lead ECG, an established risk factor for AF that reflects atrial and atrioventricular nodal conduction,^17,18^ was not associated with AF (HR 1.01, 95% CI 0.91-1.12) or with other outcomes (p > 0.19) (**Supplemental Data Table 4**). ECG-AI measures were not significantly associated with incident ischemic stroke, but they were more potent risk factors for heart failure than the imaging measure themselves. For instance, the incident heart failure HR was 1.53 for ECG-AI LAVImax (95% CI 1.30-1.81, P=3.64E-07), and only 1.36 (95% CI 1.20-1.55, P=1.23E-06) for CMR LAVImax.

**Table 1:**
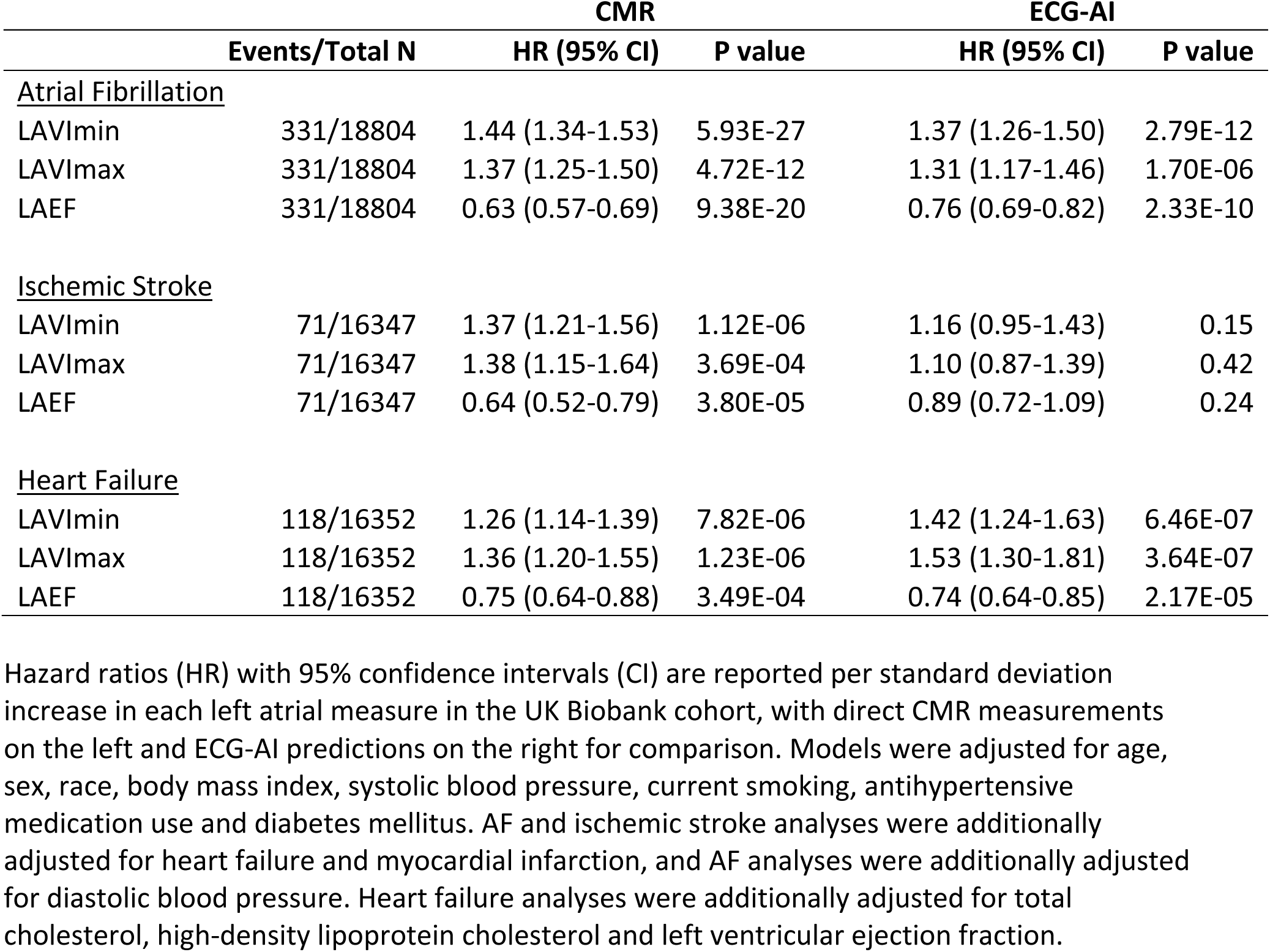
Associations of CMR and ECG-AI predicted measures of left atrial function with outcomes in the UK Biobank Study.

|  |  | CMR |  | ECG-AI |  |
| --- | --- | --- | --- | --- | --- |
|  | Events/Total N | HR (95% CI) | P value | HR (95% CI) | P value |
| <u>Atrial Fibrillation</u> |  |  |  |  |  |
| LAVImin | 331/18804 | 1.44 (1.34-1.53) | 5.93E-27 | 1.37 (1.26-1.50) | 2.79E-12 |
| LAVImax | 331/18804 | 1.37 (1.25-1.50) | 4.72E-12 | 1.31 (1.17-1.46) | 1.70E-06 |
| LAEF | 331/18804 | 0.63 (0.57-0.69) | 9.38E-20 | 0.76 (0.69-0.82) | 2.33E-10 |
| <u>Ischemic Stroke</u> |  |  |  |  |  |
| LAVImin | 71/16347 | 1.37 (1.21-1.56) | 1.12E-06 | 1.16 (0.95-1.43) | 0.15 |
| LAVImax | 71/16347 | 1.38 (1.15-1.64) | 3.69E-04 | 1.10 (0.87-1.39) | 0.42 |
| LAEF | 71/16347 | 0.64 (0.52-0.79) | 3.80E-05 | 0.89 (0.72-1.09) | 0.24 |
| <u>Heart Failure</u> |  |  |  |  |  |
| LAVImin | 118/16352 | 1.26 (1.14-1.39) | 7.82E-06 | 1.42 (1.24-1.63) | 6.46E-07 |
| LAVImax | 118/16352 | 1.36 (1.20-1.55) | 1.23E-06 | 1.53 (1.30-1.81) | 3.64E-07 |
| LAEF | 118/16352 | 0.75 (0.64-0.88) | 3.49E-04 | 0.74 (0.64-0.85) | 2.17E-05 |

Hazard ratios (HR) with 95% confidence intervals (CI) are reported per standard deviation increase in each left atrial measure in the UK Biobank cohort, with direct CMR measurements on the left and ECG-AI predictions on the right for comparison. Models were adjusted for age, sex, race, body mass index, systolic blood pressure, current smoking, antihypertensive medication use and diabetes mellitus. AF and ischemic stroke analyses were additionally adjusted for heart failure and myocardial infarction, and AF analyses were additionally adjusted for diastolic blood pressure. Heart failure analyses were additionally adjusted for total cholesterol, high-density lipoprotein cholesterol and left ventricular ejection fraction.

### Transportability of ECG-AI to external populations: MESA and CHS

To test the transportability of our findings to other populations, we applied our fully trained ECG-AI model to resting 12-lead ECGs from the baseline examinations of two cohort studies: the Multi-Ethnic Study of Atherosclerosis (MESA) and the Cardiovascular Health Study (CHS). These studies provided both age and racial diversity for validation. MESA enrolled 6,814 adults aged 45-84 years from four racial/ethnic groups (White, African American, Hispanic, and Chinese American), all free of clinical cardiovascular disease at enrollment in 2000-04. CHS is a study of 5,888 adults aged 65 and older, recruited in 1989-90 and 1992-93, with an older mean age (73 years) compared to the UKB (65 years) and MESA (62 years) (**Supplemental Data Table 2**).

We first examined the correlations between ECG-AI predictions and direct CMR measurements. While the correlations between LAVmin, LAVmax, and LAEF with ECG-AI predicted measures were lower than in UKB (Pearson’s r=0.30-0.34; **Supplemental Figure 2**), associations with cardiovascular outcomes after adjustment for established clinical risk factors were similar or greater in magnitude for each outcome.

For incident AF, risk estimates were similar across all three cohorts. Each SD higher (worse) ECG-AI predicted LAVImin was associated with greater risk: HR 1.37 (95% CI 1.26-1.50, P=2.79E-12) in the UKB test sample, HR 1.34 (95% CI 1.28-1.41, P=2.58E-30) in MESA, and HR 1.41 (95% CI 1.35-1.47, P=3.04E-49) in CHS. In contrast, each SD higher (better) ECG-AI predicted LAEF was associated with a HR of 0.76 (95% CI 0.69-0.82, P=2.33E-10) in the UKB test sample, HR 0.76 (95% CI 0.73-0.80, P=8.10E-27) in MESA, and HR 0.72 (95% CI 0.69-0.75, P=8.57E-56) in CHS (**Tables 1, 2**).

Associations of ECG-AI measures with incident ischemic stroke were less strong and similar across cohorts: HRs for LAVImin were 1.16 in the UKB test sample (95% CI 0.95-1.43, P=0.15), 1.16 in MESA (95% CI 1.03-1.30, P=0.01), and 1.22 in CHS (95% CI 1.13-1.32, P=1.88E-7). We observed the strongest associations for heart failure: each SD higher ECG-AI LAVImin was associated HRs of 1.42 in the UKB test sample (95% CI 1.24-1.63, P=6.46E-7), 1.36 in MESA (95% CI 1.23-1.50, P=4.79E-9), and 1.45 in CHS (95% CI 1.37-1.53, P=6.68E-37).

### Comparison of ECG-AI with atrial cardiopathy measures in MESA and CHS

To place these findings in context with existing, widely-studied measures of left atrial cardiopathy, we evaluated P-wave terminal force in lead V1 (PTFV1) from the 12-lead ECG and LA reservoir strain from CMR in MESA and echocardiography in CHS.^19^ For all three outcomes, ECG-AI measures outperformed PTFV1 and LA reservoir strain (**Table 2**). In MESA, LA parameters measured directly from CMR demonstrated slightly stronger associations with outcomes than their ECG-AI predicted counterparts (**Supplemental Data Table 5**).

**Table 2:**
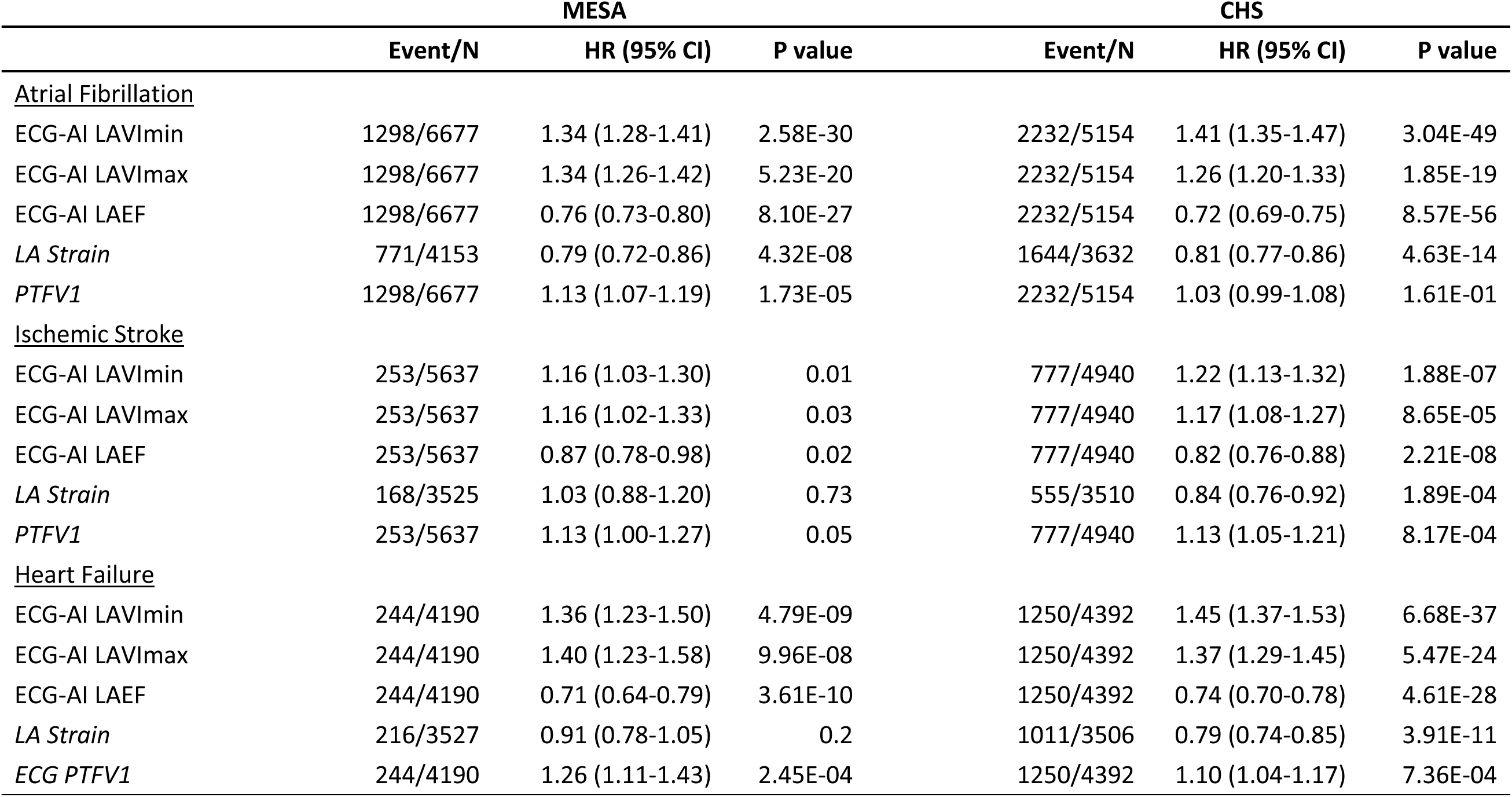
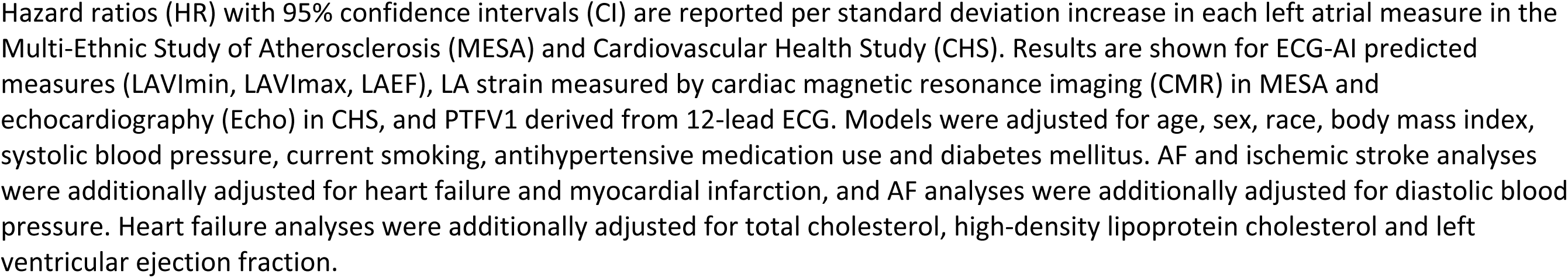
Associations of ECG-AI predicted measures and biomarkers of left atrial function with clinical outcomes in the Multi-Ethnic Study of Atherosclerosis and the Cardiovascular Health Study.

|  | MESA |  |  | CHS |  |  |
| --- | --- | --- | --- | --- | --- | --- |
|  | Event/N | HR (95% CI) | P value | Event/N | HR (95% CI) | P value |
| <u>Atrial Fibrillation</u> |  |  |  |  |  |  |
| ECG-AI LAVImin | 1298/6677 | 1.34 (1.28-1.41) | 2.58E-30 | 2232/5154 | 1.41 (1.35-1.47) | 3.04E-49 |
| ECG-AI LAVI<sub>max</sub> | 1298/6677 | 1.34 (1.26-1.42) | 5.23E-20 | 2232/5154 | 1.26 (1.20-1.33) | 1.85E-19 |
| ECG-AI LAEF | 1298/6677 | 0.76 (0.73-0.80) | 8.10E-27 | 2232/5154 | 0.72 (0.69-0.75) | 8.57E-56 |
| *LA Strain* | 771/4153 | 0.79 (0.72-0.86) | 4.32E-08 | 1644/3632 | 0.81 (0.77-0.86) | 4.63E-14 |
| *PTFV1* | 1298/6677 | 1.13 (1.07-1.19) | 1.73E-05 | 2232/5154 | 1.03 (0.99-1.08) | 1.61E-01 |
| <u>Ischemic Stroke</u> |  |  |  |  |  |  |
| ECG-AI LAVImin | 253/5637 | 1.16 (1.03-1.30) | 0.01 | 777/4940 | 1.22 (1.13-1.32) | 1.88E-07 |
| ECG-AI LAVI<sub>max</sub> | 253/5637 | 1.16 (1.02-1.33) | 0.03 | 777/4940 | 1.17 (1.08-1.27) | 8.65E-05 |
| ECG-AI LAEF | 253/5637 | 0.87 (0.78-0.98) | 0.02 | 777/4940 | 0.82 (0.76-0.88) | 2.21E-08 |
| *LA Strain* | 168/3525 | 1.03 (0.88-1.20) | 0.73 | 555/3510 | 0.84 (0.76-0.92) | 1.89E-04 |
| *PTFV1* | 253/5637 | 1.13 (1.00-1.27) | 0.05 | 777/4940 | 1.13 (1.05-1.21) | 8.17E-04 |
| <u>Heart Failure</u> |  |  |  |  |  |  |
| ECG-AI LAVImin | 244/4190 | 1.36 (1.23-1.50) | 4.79E-09 | 1250/4392 | 1.45 (1.37-1.53) | 6.68E-37 |
| ECG-AI LAVI<sub>max</sub> | 244/4190 | 1.40 (1.23-1.58) | 9.96E-08 | 1250/4392 | 1.37 (1.29-1.45) | 5.47E-24 |
| ECG-AI LAEF | 244/4190 | 0.71 (0.64-0.79) | 3.61E-10 | 1250/4392 | 0.74 (0.70-0.78) | 4.61E-28 |
| *LA Strain* | 216/3527 | 0.91 (0.78-1.05) | 0.2 | 1011/3506 | 0.79 (0.74-0.85) | 3.91E-11 |
| *ECG PTFV1* | 244/4190 | 1.26 (1.11-1.43) | 2.45E-04 | 1250/4392 | 1.10 (1.04-1.17) | 7.36E-04 |

Hazard ratios (HR) with 95% confidence intervals (CI) are reported per standard deviation increase in each left atrial measure in the Multi-Ethnic Study of Atherosclerosis (MESA) and Cardiovascular Health Study (CHS). Results are shown for ECG-AI predicted measures (LAVI<sub>min</sub>, LAVI<sub>max</sub>, LAEF), LA strain measured by cardiac magnetic resonance imaging (CMR) in MESA and echocardiography (Echo) in CHS, and PTFV1 derived from 12-lead ECG. Models were adjusted for age, sex, race, body mass index, systolic blood pressure, current smoking, antihypertensive medication use and diabetes mellitus. AF and ischemic stroke analyses were additionally adjusted for heart failure and myocardial infarction, and AF analyses were additionally adjusted for diastolic blood pressure. Heart failure analyses were additionally adjusted for total cholesterol, high-density lipoprotein cholesterol and left ventricular ejection fraction.

In, MESA, we also evaluated correlations between LA parameters with other ECG and imaging measures of cardiac structure and function. The correlations of both the ECG-AI predicted and CMR-measured LA parameters with left ventricular ejection fraction (LVEF) and with PTFV1 were low, with all Pearson correlations below |0.2| (**Supplemental Data Table 6**). Correlations with log NT-proBNP, a sensitive but nonspecific protein biomarker of ventricular stretch associated with several cardiovascular outcomes, were stronger for ECG-AI measures (|r| = 0.33-0.37) than for CMR measures (|r| = 0.16-0.28).

Given that the widely available clinical biomarker NT-proBNP is a strong predictor of AF, cardioembolic stroke, and heart failure, and is moderately correlated with LA ECG-AI measures, we additionally adjusted for log NT-proBNP. Associations between ECG-AI measures adjusted for log NT-proBNP with AF, ischemic stroke, and heart failure were attenuated, but remained significantly associated with outcomes (**Supplemental Data Table 8**).

### Ischemic stroke subtypes

Most ischemic strokes can be classified by their likely causative mechanisms based on neuroimaging and clinical context. Large-artery atherosclerotic (previously called atherothrombotic), small vessel (i.e., lacunar strokes), and cardioembolic are the most common etiologies^20^. ECG-AI measures were strongly associated with cardioembolic stroke, the hallmark complication of both AF and atrial cardiopathy^21^, but not large-artery atherosclerotic or small-vessel strokes, as hypothesized. For example, each SD higher ECG-AI LAVImin was associated with a 38% greater risk of cardioembolic stroke in MESA (95% CI 14-69%, P=1.27E-03) and 66% greater risk in CHS (95% CI 47-86%, P=3.67E-17), while each SD higher ECG-AI LAEF was associated with a 30% decreased risk of cardioembolic stroke in MESA (95% CI 16-42%, P=2.01E-04) and a 41% decreased risk in CHS (95% CI 34-47%, P=1.49E-20) (**Table 3**). Conversely, none of the ECG-AI measures were associated with small vessel or large artery atherosclerotic stroke (P >= 0.37).

**Table 3:**
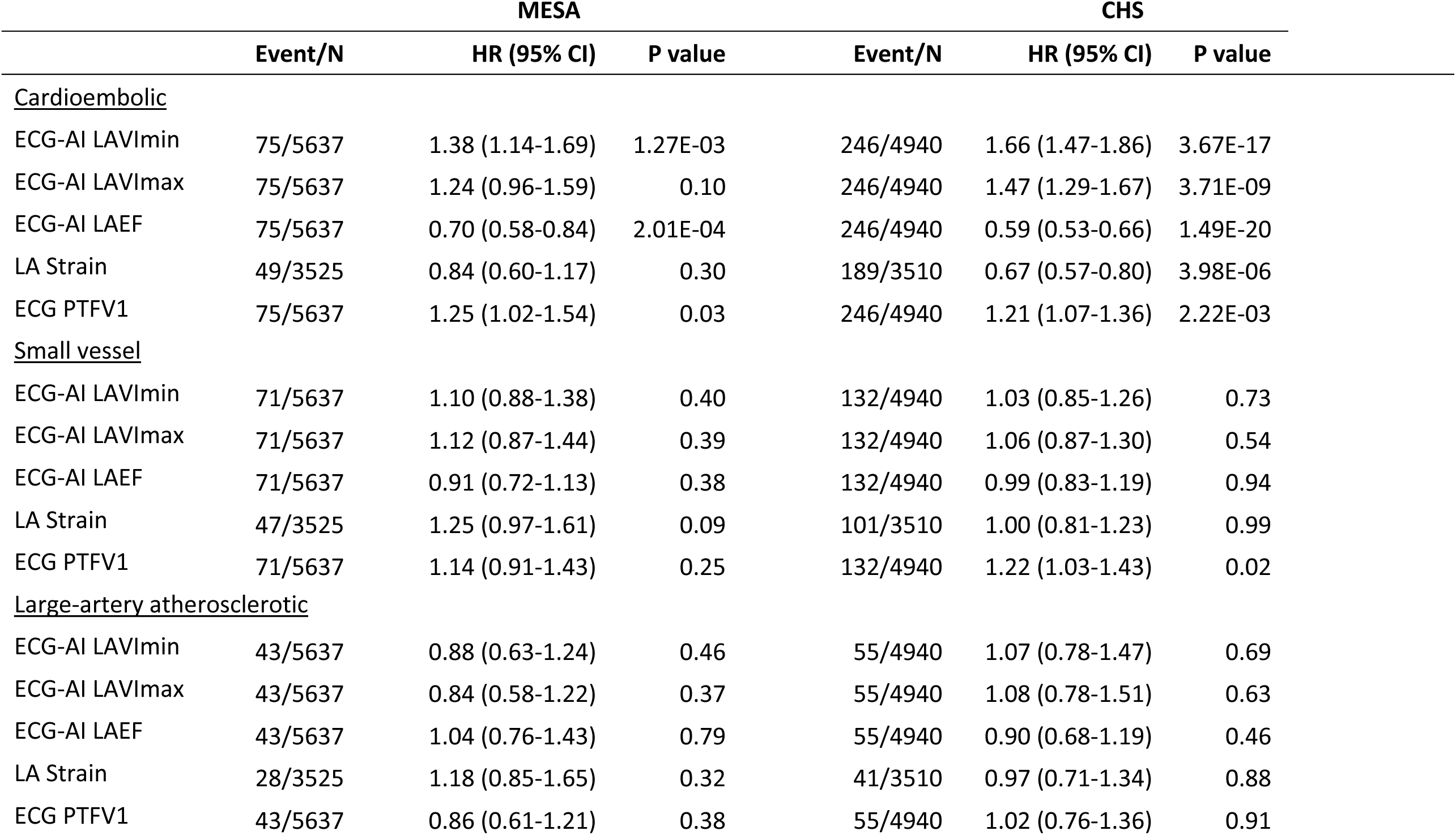
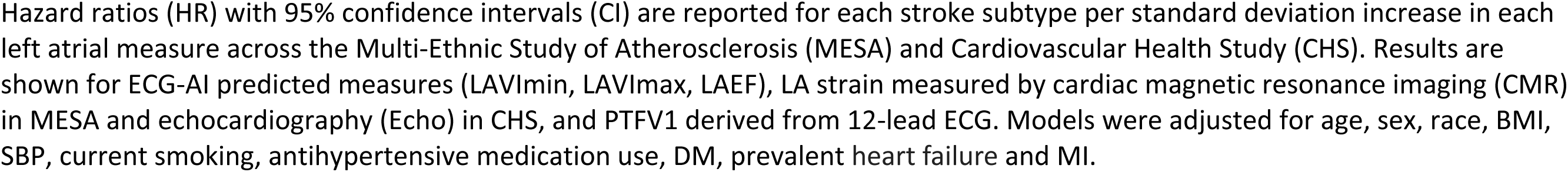
Associations of left atrial function measures with stroke subtypes in the Multi-Ethnic Study of Atherosclerosis and the Cardiovascular Health Study.

|  | MESA |  |  | CHS |  |  |
| --- | --- | --- | --- | --- | --- | --- |
|  | Event/N | HR (95% CI) | P value | Event/N | HR (95% CI) | P value |
| <u>Cardioembolic</u> |  |  |  |  |  |  |
| ECG-AI LAVImin | 75/5637 | 1.38 (1.14-1.69) | 1.27E-03 | 246/4940 | 1.66 (1.47-1.86) | 3.67E-17 |
| ECG-AI LAVImax | 75/5637 | 1.24 (0.96-1.59) | 0.10 | 246/4940 | 1.47 (1.29-1.67) | 3.71E-09 |
| ECG-AI LAEF | 75/5637 | 0.70 (0.58-0.84) | 2.01E-04 | 246/4940 | 0.59 (0.53-0.66) | 1.49E-20 |
| LA Strain | 49/3525 | 0.84 (0.60-1.17) | 0.30 | 189/3510 | 0.67 (0.57-0.80) | 3.98E-06 |
| ECG PTFV1 | 75/5637 | 1.25 (1.02-1.54) | 0.03 | 246/4940 | 1.21 (1.07-1.36) | 2.22E-03 |
| <u>Small vessel</u> |  |  |  |  |  |  |
| ECG-AI LAVImin | 71/5637 | 1.10 (0.88-1.38) | 0.40 | 132/4940 | 1.03 (0.85-1.26) | 0.73 |
| ECG-AI LAVImax | 71/5637 | 1.12 (0.87-1.44) | 0.39 | 132/4940 | 1.06 (0.87-1.30) | 0.54 |
| ECG-AI LAEF | 71/5637 | 0.91 (0.72-1.13) | 0.38 | 132/4940 | 0.99 (0.83-1.19) | 0.94 |
| LA Strain | 47/3525 | 1.25 (0.97-1.61) | 0.09 | 101/3510 | 1.00 (0.81-1.23) | 0.99 |
| ECG PTFV1 | 71/5637 | 1.14 (0.91-1.43) | 0.25 | 132/4940 | 1.22 (1.03-1.43) | 0.02 |
| <u>Large-artery atherosclerotic</u> |  |  |  |  |  |  |
| ECG-AI LAVImin | 43/5637 | 0.88 (0.63-1.24) | 0.46 | 55/4940 | 1.07 (0.78-1.47) | 0.69 |
| ECG-AI LAVImax | 43/5637 | 0.84 (0.58-1.22) | 0.37 | 55/4940 | 1.08 (0.78-1.51) | 0.63 |
| ECG-AI LAEF | 43/5637 | 1.04 (0.76-1.43) | 0.79 | 55/4940 | 0.90 (0.68-1.19) | 0.46 |
| LA Strain | 28/3525 | 1.18 (0.85-1.65) | 0.32 | 41/3510 | 0.97 (0.71-1.34) | 0.88 |
| ECG PTFV1 | 43/5637 | 0.86 (0.61-1.21) | 0.38 | 55/4940 | 1.02 (0.76-1.36) | 0.91 |

Hazard ratios (HR) with 95% confidence intervals (CI) are reported for each stroke subtype per standard deviation increase in each left atrial measure across the Multi-Ethnic Study of Atherosclerosis (MESA) and Cardiovascular Health Study (CHS). Results are shown for ECG-AI predicted measures (LAVI<sub>min</sub>, LAVI<sub>max</sub>, LAEF), LA strain measured by cardiac magnetic resonance imaging (CMR) in MESA and echocardiography (Echo) in CHS, and PTFV1 derived from 12-lead ECG. Models were adjusted for age, sex, race, BMI, SBP, current smoking, antihypertensive medication use, DM, prevalent heart failure and MI.

### Prediction evaluation

We developed 5-year prediction models for each outcome, comparing four approaches: (1) base model (age and sex only) (2) base model and a weighted linear combination of the three ECG-AI measures, (3) CHARGE-AF clinical risk score, and (4) CHARGE-AF and ECG-AI measures. Model weights were estimated in UKB and externally validated in MESA and CHS. There were 842 AF events and 63 cardioembolic strokes across MESA and CHS.

For 5-year AF prediction (**Figure 2, Supplemental Table 9**), the ECG-AI model performed similarly to the CHARGE-AF model in terms of area under the received operating characteristic curve (AUROC, 0.75 vs 0.74) but had a substantially better area under the precision recall curve (AUPRC, 0.23 vs 0.18). Moreover, the addition of ECG-AI to the CHARGE-AF improved AUROC (Δ AUROC 0.024, 95% CI 0.017-0.034, P < 10E-4) and AUPRC (Δ AUPRC 0.046, 95% CI 0.030-0.063, P < 10E-4). For the prediction of cardioembolic stroke, ECG-AI outperformed CHARGE-AF in AUROC (0.82 vs 0.80) and AUPRC (0.033 vs 0.026), and the addition of ECG-AI to CHARGE-AF improved prediction (Δ AUROC 0.037, 95% CI 0.008-0.066, P=0.012). When categorizing risk predictions into tertiles, the high-risk ECG-AI group has a OR of 52.8 (95% CI 7.3-382, P = 8.5E-5) compared to the low-risk group.

**Figure 2:**
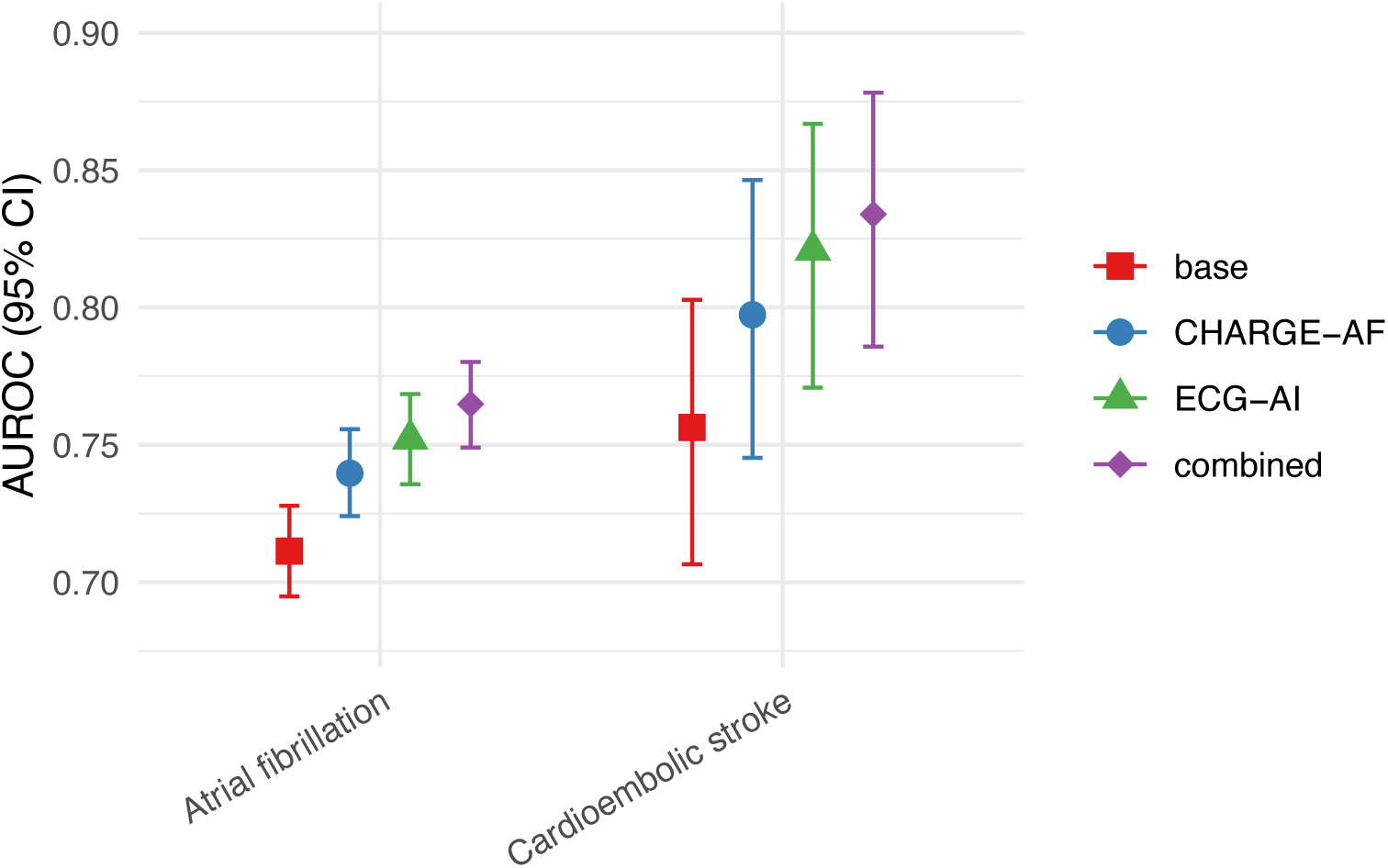
Classification performance of 5-year AF risk scores. Receiver operating characteristic area under the curve (AUC) for 5-year AF risk score classification performance in combined MESA and CHS cohorts. The prediction models compared are: (1) age and sex only (base), (2) base + ECG-AI left atrial measures (ECG-AI), (3) CHARGE-AF risk score alone (CHARGE-AF), and (4) CHARGE-AF + ECG-AI (combined). Models were evaluated for 5-year risk of AF and cardioembolic stroke. Plots show AUC values with 95% confidence intervals.

### Subclinical AF screening

AF can be difficult to detect because it is paroxysmal and asymptomatic in many individuals. One potential application for a tool that estimates LA function is the selection of high-risk individuals who might benefit from AF screening with extended cardiac monitoring, a sensitive and unbiased method of arrhythmia detection^22^. In MESA, a resting 12-lead ECG was performed at Exam 5 (2010-12) and 14-day ambulatory cardiac monitoring was performed an average of six years later at Exam 6 (2016-18). We applied our ECG-AI model to Exam 5 ECGs and used the highest quintile of predicted LAVImin or LAVImax to identify a high-risk population. Among the 20% of participants identified as high-risk, 14% had AF on cardiac monitoring (positive predictive value, PPV), and 47% of all cases of monitor-detected AF were identified (sensitivity). By comparison, among high-risk individuals within the highest quintile of a widely used risk prediction tool, CHARGE-AF^23^, for AF, the PPV was 11% and the sensitivity was 36%. The AUCs for the high-risk quintile predicting monitor-detected AF were 0.64 (95% CI 0.59-0.70) for ECG-AI LAVImin and 0.59 (95% CI 0.53-0.64) for CHARGE-AF (**Supplemental Data Table 10**). When comparing high- vs low-risk tertiles, the relative risk for subclinical AF with CHARGE-AF was 6.0 (95% CI 2.9-12.3), and for ECG-AI it was 8.2 (95% CI 3.7-18.3).

### Gradient-weighted Class Activation Mapping

To identify which ECG features most influenced our model predictions, we analyzed Gradient-weighted Class Activation Mapping (Grad-CAM) intensities for the 50 highest-risk ECGs (those predicting the largest LA volumes and lowest LAEF values) (**Figure 1, Supplemental Figure 3**). For predicting LAVmin volumes, the model primarily focused on the second half of the P wave, with a secondary focus on the R peak and the ascending portion of the T wave. For LAVmax predictions, the model was broadly focused on the entire P-waveform, with an emphasis on the first half of the P wave. When predicting lower LAEF, the influential regions spanned more broadly across the P-QRS waveforms but were most pronounced over the PQ segment. Notably, these Grad-CAM intensity patterns were consistent across all three cohort studies. These findings suggest that our model exhibits robust and generalizable feature identification.

### Genetic analyses of ECG-AI phenotypes

To uncover some of the biological factors driving our ECG-AI measures, we used human genomic data. We performed genome-wide association study (GWAS) analyses of five ECG-AI measures (indexed and non-indexed volumes and LAEF) among European ancestry participants in the UK Biobank Study test sample who were free of AF at baseline (N=33,245) (**Supplemental Figure 1**). We identified 12 loci across the five traits (**Supplemental Data Table 11, Supplemental Figure 4**). The strongest association was observed for the T allele of rs11153730 (coded allele frequency 0.51, Beta=-0.085, SE=0.008, P=8.1E-27), a well-characterized variant near *PLN* that is a locus for multiple ECG metrics, heart failure, and AF. The estimated heritability (*h²*) of ECG-AI traits was modest, ranging from 0.14 for LAVmax to 0.17 for LAVImax (**Supplemental Data Table 12**). Notably, half of these loci do not have genomic targets that overlap with AF loci^24^.

Using these GWAS results, we evaluated genetic correlations between ECG-AI predictions, CMR measurements, and established cardiovascular risk factors. ECG-AI measures demonstrated moderate genetic correlation with their CMR counterparts: h² of 0.47 for LAVImin, 0.34 for LAVImax, and 0.62 for LAEF. We found left ventricular mass (LVM) indexed to BSA had a strong positive genetic correlation with ECG-AI volume measures and was negatively correlated with LAEF (**Figure 3**). LAEF was also genetically correlated with AF and heart failure, but the volume measures were not. The ECG-AI volume measures exhibited genetic correlations with type 2 diabetes and body mass index that were not present with the directly measured CMR volume traits.

**Figure 3:**
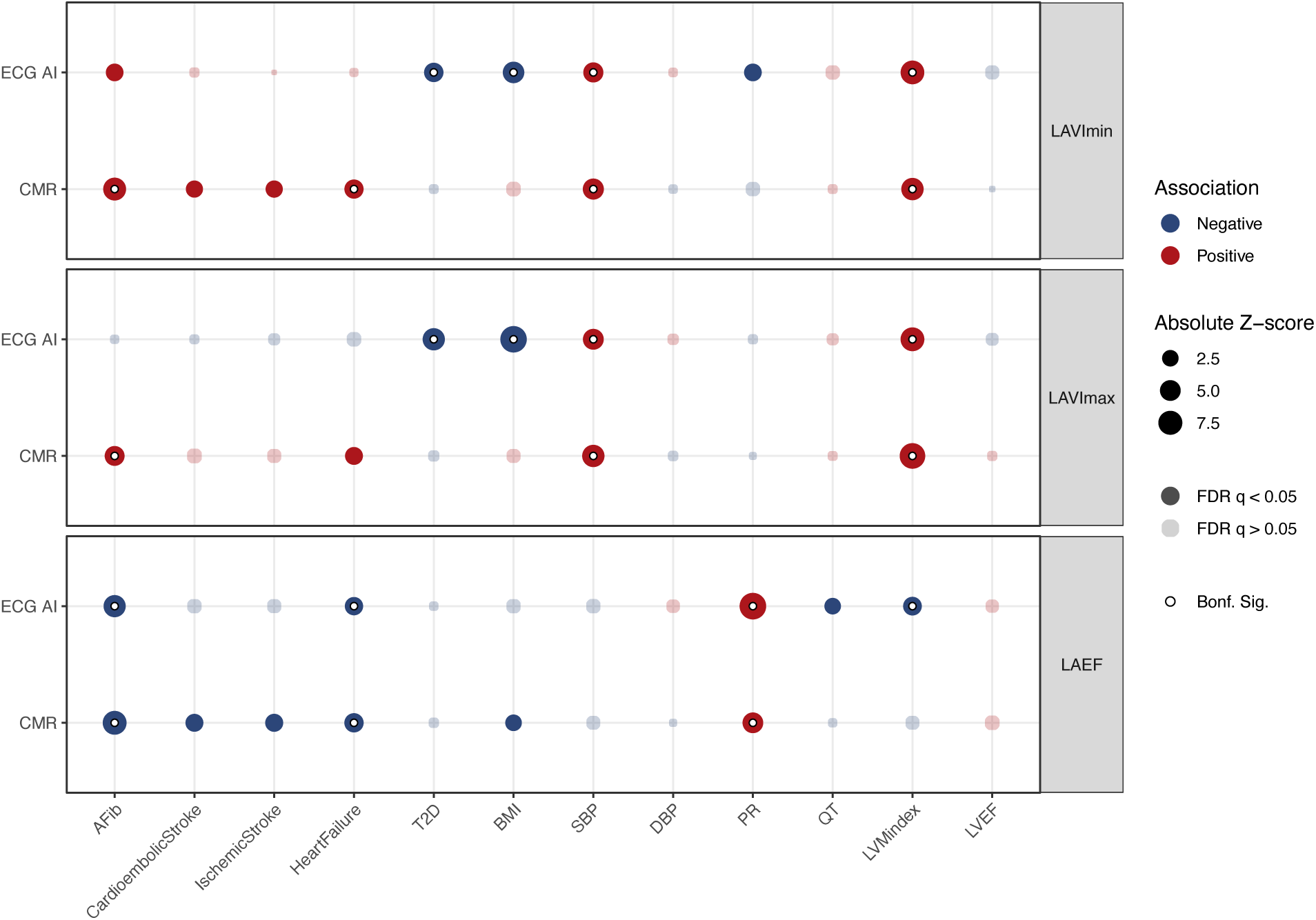
Genetic Correlation of ECG-AI and CMR atrial measures with cardiovascular traits. LD score regression shows the genetic correlation between ECG-AI prediction measures, directly measured CMR traits, and cardiovascular traits. Each panel corresponds to a different left atrial parameter left atrial ejection fraction (LAEF), left atrial maximal volume indexed to BSA (LAVImax), and left atrial minimal volume indexed to BSA (LAVImin). The size of the bubble represents the absolute Z-score for each trait. Positive cross-trait associations are in red, while negative associations are in blue. Associations with a significant correlation after Bonferroni correction contain a white core, and those with a significant correlation after controlling for the false discovery rate (FDR) are saturated (dark), while insignificant correlations are in greyscale.

## Discussion

We have demonstrated that deep learning models trained on high-quality cardiac magnetic resonance imaging can predict LA function from the resting 12-lead ECG, an inexpensive and widely available test. Our ECG-AI measures were strongly associated with incident AF, incident heart failure, and incident ischemic stroke, independent of established risk factors and biomarkers for these outcomes, with a remarkable specificity for the cardioembolic stroke subtype. That our ECG-AI measures outperformed conventional ECG and imaging measures of atrial cardiopathy suggests that our deep learning model extracts novel information from cardiac imaging data. Human genetic evidence suggests that despite some overlap in genomic loci and modest genetic correlation with AF, the ECG-AI measures may reflect different underlying mechanisms of cardiovascular risk. The consistent performance of these ECG-AI measures in two external cohort studies without additional model training or refinement demonstrates the reproducibility and transportability of our findings across diverse populations. Moreover, compared to a clinical risk prediction tool, our ECG-AI improved the 5-year prediction of AF and cardioembolic stroke, the hallmark complication of AF, and identified nearly half of all cases of subclinical AF in the highest risk quintile, a group who may benefit from screening.

Previous deep learning approaches applied to ECG analysis have primarily focused on predicting the risk of clinically identified outcomes. For instance, Attia et al.^13^ demonstrated that AI-enabled ECG could identify AF from ECGs in sinus rhythm after or within 31 days of diagnosis, with an AUC of 0.87. Similarly, Raghunath et al.^25^ developed an ECG-AI model to detect paroxysmal AF with comparable performance. These classification models effectively served as “black boxes,” identifying patterns associated with AF without explicitly modeling the underlying pathophysiological changes. There have been numerous highly capable ECG models developed for arrhythmia detection^26^, with some achieving physician-level performance but providing limited insight into structural cardiac abnormalities preceding arrhythmogenesis and biological mechanisms.^27^ These models are inherently limited by their reliance on a clinical diagnosis of AF, which is present but undetected in a large proportion of individuals at a rate that varies by race.^15,28^

In contrast to these binary classification approaches, our research represents a fundamental shift toward quantitative prediction of continuous LA parameters that reflect the substrate for AF development. While previous models asked, “Does this patient have AF?” our model asks, “What is the state of this patient’s left atrium?” This latter approach may offer advantages. First, by directly modeling LA volumes and function—established imaging biomarkers with known pathophysiological relevance—our predictions provide mechanistic insights into the atrial substrate. Second, our model outputs continuous estimates of cardiac structure and function, potentially enabling more granular risk stratification than binary classification. Third, by directly predicting atrial volume and function, we avoid the ascertainment bias inherent in models that rely on clinically detected AF. Fourth, the moderate genetic correlation between our ECG-AI and CMR measures (*h*² of 0.47-0.62), coupled with identification of genetic loci shared with AF, provides biological validation lacking in previous classification models. Lastly, atrial cardiopathy itself is a target for disease prevention and clinical trials of therapeutics are already underway^29,30,31^; an inexpensive and widely available tool for the detection of atrial cardiopathy will accelerate the development of new therapeutic approaches to reduce atrial cardiopathy-related complications.

A notable finding was the specificity of ECG-AI measures for cardioembolic stroke and not for other stroke subtypes. This specificity aligns with the pathophysiological mechanisms linking atrial dysfunction to thromboembolism. Stroke subtypes were rigorously adjudicated in the cohort studies, which made this comparison possible. Our findings support the use of ECG-AI measures in risk-stratification for cardioembolic stroke, a condition that often requires different preventive strategies than other stroke subtypes.

Our Grad-CAM analyses revealed additional mechanistic insight on the ECG features most informative for predicting the left atrial measures, with the ECG-AI model focused on different regions of the cardiac cycle for each left atrial measure. The LAVmin model was focused on the second half of the P wave for LAVmin, while LAVmax model was focused on the first half. In contrast, the LAEF signal was focused during atrioventricular repolarization on the PQ segment, suggesting that our model captures physiological signals consistent with our understanding of cardiac electrophysiology.

Genetic analyses further characterized the biological basis of our ECG-AI predictions. Of the 12 genetic loci we discovered, half were located within 500 kb of known AF variants, supporting the biological relevance of our ECG-AI measures. The moderate genetic correlations between ECG-AI and CMR measures (*h²* of 0.47 for LAVImin, 0.34 for LAVImax, and 0.62 for LAEF) suggest that our model captures a substantial portion of the heritable component of left atrial structure and function. However, the genetic correlations between our ECG-AI measures and related risk factors and outcomes differed from directly measured CMR variables.

As in previous studies^32,33^, we found plasma protein NT-proBNP was a potent risk marker for AF and ischemic stroke, and our ECG-AI measures were significantly associated even after adjusting for this sensitive biomarker of cardiac stretch that is widely performed clinically. This relationship suggests that our ECG-AI approach provides additional prognostic information beyond that captured by this established biomarker, and that combining these complementary biomarkers can improve the identification of individuals at elevated cardiovascular risk.

Our ECG-AI prediction model performed as well as the most widely used clinical risk prediction tool for AF in 5-year prediction analyses for predicting AF and cardioembolic stroke, and enhanced prediction when combined for AF. For identifying high-risk individuals to undergo AF screening with extended cardiac monitoring, perhaps the most important clinical application, ECG-AI measures outperformed the clinical prediction tool and suggest an urgent need to evaluate targeted screening approach for AF based on cardiac function estimated with ECG-AI. Performing 14-day cardiac monitoring shortly after screening with a 12-lead ECG is likely to result in far superior performance compared with our analyses in MESA, where cardiac monitoring followed the 12-lead ECG by an average of six years. While cardiac imaging remains the gold standard for assessing left atrial function, its widespread application is limited by high costs, availability, and the need for expert interpretation. Our findings demonstrate that AI-enhanced ECG analysis can extract clinically meaningful information from widely available and inexpensive 12-lead ECGs, yielding remarkably similar associations to clinical outcomes as the imaging-derived measurements.

This study has several limitations. Our ECG-AI model was trained on LA volumes predicted by a segmentation model, which, despite quality control, may contain errors. The correlations between ECG-AI and CMR-derived measures were only moderate (0.40-0.50). The UKB population comprises predominantly European ancestry individuals who are healthier than the general population. The validation of our ECG-AI in a large multi-ethnic cohort study demonstrates the tremendous potential for transportability of these findings and models to diverse and heterogeneous populations.^34,35^ Our analysis of subclinical AF included only 86 detected cases and should be replicated in larger studies. Although we demonstrated external validation in two independent research cohorts, broader validation across diverse clinical settings and ECG acquisition protocols is needed to establish widespread clinical utility.

Our ECG-AI approach represents a promising, accessible tool for assessing left atrial function and stratifying cardiovascular risk using standard 12-lead ECGs. In contrast with more resource- and technology-intensive methods such as cardiac imaging, this approach could democratize improve access to advanced cardiovascular risk assessment across diverse healthcare settings. The public availability of our model facilitates further validation and implementation studies, potentially accelerating translation into clinical practice.

## Acknowledgements

*UK Bioban*k: This research has been conducted using the UK Biobank Resource under Application Number #40713. The authors would like to thank the participants and researchers involved in creating and maintaining the UK Biobank as a valuable resource for health-related research.

*Multi-Ethnic Study of Atherosclerosis:* This research was supported by contracts 75N92020D00001, HHSN268201500003I, N01-HC-95159, 75N92020D00005, N01-HC-95160, 75N92020D00002, N01-HC-95161, 75N92020D00003, N01-HC-95162, 75N92020D00006, N01-HC-95163, 75N92020D00004, N01-HC-95164, 75N92020D00007, N01-HC-95165, N01-HC-95166, N01-HC-95167, N01-HC-95168 and N01-HC-95169 from the National Heart, Lung, and Blood Institute, grants UL1-TR-000040 and UL1-TR-001420 from NCATS, and UL1-RR-025005 from NCRR and R01-HL-127659, R01-HL-107577 and R01 HL127659 from NHLBI. The authors thank the other investigators, the staff, and the participants of the MESA study for their valuable contributions. A full list of participating MESA investigators and institutions can be found at http://www.mesa-nhlbi.org.

*Cardiovascular Health Study:* This Cardiovascular Health Study research was supported by NHLBI contracts HHSN268201200036C, HHSN268200800007C, HHSN268201800001C, N01HC55222, N01HC85079, N01HC85080, N01HC85081, N01HC85082, N01HC85083, N01HC85086, N01HC85084, N01HC35129, R01AG15928, R01AG20098, 75N92021D00006; and NHLBI grants U01HL080295, R01HL087652, R01HL103612, R01HL105756, R01HL120393, U01HL130114, and R01HL172803 with additional contribution from the National Institute of Neurological Disorders and Stroke (NINDS). Additional support was provided through R01AG023629 from the National Institute on Aging (NIA). A full list of principal CHS investigators and institutions can be found at CHS-NHLBI.org. The content is solely the responsibility of the authors and does not necessarily represent the official views of the National Institutes of Health.

JSF was supported by NIH grants HL142599, HL149706, and HL172495.

## Data availability

UK Biobank data are made available to researchers from research institutions as described at this site: https://www.ukbiobank.ac.uk/enable-your-research. Data for Multi-Ethnic Study of Atherosclerosis and Cardiovascular Health Study can be requested at https://internal.mesa-nhlbi.org/ and https://chs-nhlbi.org/node/6222, respectively. GWAS summary statistics are publicly available under accession numbers GCST006414 (atrial fibrillation), GCST90104540 (ischemic stroke), GCST90104541 (cardioembolic stroke), GCST90104542 (large artery stroke), GCST90104543 (small vessel stroke), GCST90132314 (coronary artery disease), GCST90239658 (low density lipoprotein cholesterol), GCST90239652 (high density lipoprotein cholesterol), GCST90239664 (triglycerides), GCST90310294 (systolic blood pressure), GCST90310295 (diastolic blood pressure), GCST009004 (BMI) and GCST010320 (PR interval) at https://www.ebi.ac.uk/gwas/home. The GWAS summary statistics for heart failure (https://cvd.hugeamp.org/dinspector.html?dataset=Shah2020_HF_EU), type 2 diabetes (https://www.diagram-consortium.org/downloads.html), and QT interval (https://arkinglab.org/upload/QT/) are available at their respective repositories.

## Code availability

The fully trained model and weights are available for download at https://github.com/j-brody/ecg_ai_atrialmeasures.

## Author contributions

J.A.B and J.S.F. designed the study. J.A.B., C.M.S and K.L.W. performed analyses. W.L., J.A.L., E.Z.S., S.R.H. contributed data from MESA. B.M.P, E.Z.S., S.R.H., W.L. contributed data from CHS. J.A.B, V.Y and J.S.F. drafted the manuscript. All authors reviewed the manuscript and provided critical revision.

## Online Methods

### UK Biobank

The UK Biobank (UKB) is a cohort study that recruited 500,000 participants aged 40 to 69 years from the United Kingdom between 2006 and 2010^36^. The baseline assessment included comprehensive questionnaires, blood and urine collection for biochemical measurements, anthropometry, and interviews. All UKB participants underwent whole-genome genotyping through the UK Biobank Whole-Genome Sequencing Consortium^37^.

All participants provided electronic informed consent at the UK Biobank assessment centers. The UKB received ethics approval from the North West Multi-Centre Research Ethics Committee.

Longitudinal follow-up for health outcomes of atrial fibrillation (AF), stroke, and heart failure was performed through linkages with national databases on hospitalizations, primary care records, and death registries. Incident AF events were identified from self-reports, nurse interviews, hospital admission codes, death register cause-of-death codes, and UK Biobank first occurrence data, while ischemic stroke and heart failure were identified solely from diagnosis codes and cause-of-death codes (**Supplemental Data Table 13**). Participants diagnosed with incident AF before the imaging visit or those with pacemakers were excluded from all analyses. Additionally, participants whose imaging visits occurred after the UKB data censoring dates were excluded. Follow-up for this study extended through 2022. Hospital data were available until October 2022 for England, August 2022 for Scotland, and May 2022 for Wales. Death record data were censored in November 2022 across all sites.

### Multi-Ethnic Study of Atherosclerosis

The Multi-Ethnic Study of Atherosclerosis (MESA) is a community-based observational study of 6814 men and women, free of known CVD at baseline, representing four racial/ethnic groups (white, African American, Hispanic, and Chinese-American), aged 45–84 years and free of clinical CVD at enrollment^38^. Study participants were recruited in 2000-2 at 6 field centers in the US (Baltimore, MD; Chicago, IL; Forsyth County, NC; Los Angeles, CA; New York, NY; and St. Paul, MN). At telephone contacts every 9–12 months during follow-up, participants were asked to identify new hospitalizations and diagnoses, and medical records were obtained. Institutional review boards of all field centers approved the study protocol, and all participants gave written informed consent.

Clinically-recognized AF was identified by an International Classification of Disease (ICD) hospital discharge diagnosis code (version 9: 427.31 or 427.32; version 10: I48) in any position; and for those enrolled in fee-for-service Medicare, by an inpatient, outpatient, or physician claim with an AF code^39^.

A panel reviewed medical records and adjudicated heart failure events according to standardized criteria. The present study included probable or definite heart failure events. Probable heart failure required heart failure symptoms, a physician diagnosis of heart failure, and treatment. Definite heart failure required at least one objective feature of heart failure, including abnormalities on chest X-ray, echocardiography, or ventriculography.

Methods for clinical ischemic stroke adjudication in MESA has been previously described^40^. In brief, information about all new cardiovascular conditions, hospital admissions, cardiovascular outpatient diagnoses, treatments, and deaths were obtained. Two vascular neurologists from the MESA study events committee independently reviewed all medical records for end point classification and assignment of incidence dates. They reviewed and classified stroke as present if there was a focal neurologic deficit lasting 24 hours or until death or, if <24h, there was a clinically relevant lesion on brain imaging and there was no nonvascular cause. Patients with focal neurological deficits secondary to brain trauma, tumor, infection, or other nonvascular cause were excluded. Ischemic strokes were distinguished from hemorrhagic stroke using findings on imaging, surgery, autopsy, or some combination of these. Ischemic stroke subtypes were assigned based on an extension of the Trial of Org 10172 in Acute Stroke Treatment (TOAST) criteria to try to reduce the number classified as undetermined.

### Cardiovascular Health Study

The Cardiovascular Health Study (CHS) is a cohort study of risk factors for cardiovascular diseases in adults aged 65 years and older, recruited from four field centers in the United States (Sacramento, CA; Hagerstown, MD; Winston-Salem, NC; Pittsburgh, PA)^41^. Between June 1989 and June 1990, 5,201 participants were recruited from random samples of people from Medicare eligibility lists, and an additional 687 predominantly Black participants were recruited between November 1992 and June 1993. Study participants were seen in the clinic annually between enrollment and 1998-99 and were contacted by telephone at 6-month intervals through June 2015 to collect information to ascertain changes in health status, hospitalizations, and medication use^42^. Institutional review boards of all participating sites approved the study, and all study participants provided informed consent.

AF events were identified using a combination of electrocardiographic (ECG) findings from clinic visits through 1999, hospital discharge records indicating AF, and Medicare inpatient, outpatient, and physician (carrier) claims data. Resting 12-lead ECGs were performed at baseline and follow-up visits, with centralized readings to classify AF according to standardized criteria. For AF identified through Medicare data, diagnosis required either ECG evidence, one inpatient claim, or two outpatient or physician claims within 365 days using ICD-9-CM codes 427.31 or 427.32 in any diagnostic position.

Assessment of heart failure events in CHS has been previously described^43^. All potential heart failure events were identified through in-person visits or telephone interviews. These events were then adjudicated by the central events committee. Adjudication of heart failure was based on a physician diagnosis of heart failure, documented symptoms and signs of heart failure, use of medical treatment for heart failure, and supportive findings on diagnostic testing.

Methods for clinical ischemic stroke adjudication in CHS has been previously described^44^. In summary, an expert events committee, including neurologists from the study sites and CHS coordinating center, along with a neuroradiologist from the imaging reading center, reviewed the cases. Ischemic stroke was characterized by the sudden onset of a focal neurological deficit persisting for at least 24 hours, supported by compatible imaging findings on computed tomography scan or cardiac magnetic resonance imaging (CMR), and without evidence of intracranial hemorrhage or a nonvascular underlying cause. The adjudication committee categorized ischemic strokes into four subtypes based on the presumed infarction mechanism: cardioembolic, small vessel, large-artery atherosclerotic, or unknown/other causes.

### Cardiac magnetic resonance imaging parameters

The UK Biobank Imaging Study began in 2014 and included magnetic resonance imaging of the brain, abdomen, and heart^45^. The cardiac magnetic resonance imaging (CMR) protocol consists of a 20-minute scan performed using a 1.5 Tesla scanner (MAGNETOM Aera, Syngo Platform VD13A, Siemens Healthcare, Erlangen, Germany)^46^. Cardiac volume and function parameters were obtained through three long-axis cines and complex short-axis stacks of balanced, steady-state free precession cines^46^. The first 5,065 CMR scans were manually analyzed, and LA contours at end-systole and end-diastole were traced^47^. Using these manually annotated images, Bai et al.^16^ developed a fully automated convolutional network pipeline for the large-scale derivation of measures of cardiac structure and function. The accuracy of these automated methods was evaluated quantitatively by various technical metrics (Dice metric, contour distance metrics, and Hausdorff distance) and qualitatively by clinical experts^16^. This algorithm is publicly available to the research community through GitHub.

We applied this fully convolutional network to all CMR scans from the first imaging visit available as of October 8th, 2023, to obtain left atrial minimal volume and left atrial maximal volume. Images from the first visit CMR studies were acquired from April 2014 to May 2023. Left atrial volumes were indexed to body surface area to obtain left atrial indexed minimal volume (LAVImin) and left atrial indexed maximal volume (LAVImax). Left atrial ejection fraction (LAEF) was calculated through the following formula: [(maximal volume – minimal volume)/maximal volume *(100)]. We then manually reviewed images when the estimated measurements were biologically implausible and identified threshold values for each parameter to exclude these measurements from further analyses.

In MESA, CMR imaging was performed at Exam 1 (2000-04, n=5,004) and Exam 5 (2011-13, n=3,016). Long-axis cine images acquired during a 30–45-minute protocol using 1.5-T scanners with ECG-gated fast gradient echo pulse sequence at baseline and steady-state free precession at Exam 5^48,49^. Ventricular measures were obtained by manually tracing the endocardial and epicardial borders of short-axis cine images at end-systole and end-diastole^50^. LA strain and phasic volume measures were obtained separately from baseline 2- and 4-chamber cine images. LA endocardial and epicardial borders were manually defined at end-systole by an experienced operator using Multimodality tissue tracking software (MTT, Toshiba, Tokyo, Japan). LA maximum, pre-atrial, and minimum contraction volumes were extracted from volume curves generated using area-length methods. The MTT software generated longitudinal strain curves for each atrial wall segment, and global longitudinal strain was calculated by averaging all segmental values at each time frame. The peak (reservoir) longitudinal LA strain value was measured from the global longitudinal strain curve^51^.

### Electrocardiograms

In UKB, ECG waveforms were extracted for all 62,926 subjects with resting digital ECGs measured at the Instance 2 visit that were available for download as of Oct 8th, 2023. The median beat waveform from ECGs sampled at 500 Hz in each lead were extracted as Extensible Markup Language files. We included eight non-derived leads from the 12-lead ECG: leads I, II, and V1 to V6. The remaining leads are mathematical transformations and do not include additional information. ECGs with automated diagnosis of poor quality, suspect arm reversal, undetermined rhythm or implausible ventricular rates of <20 or >140 were excluded. Additionally, ECGs with atrial fibrillation, atrial flutter or pacemaker were excluded prior to model training or analysis.

In CHS and MESA, resting 10-second 12-lead ECGs sampled at 500 Hz were recorded in a uniform fashion in all participants at baseline using MAC PC ECG Machines (Marquette Electronics Inc, Milwaukee, WI). All ECGs were visually inspected for technical errors and inadequate quality centrally at the Epidemiological Cardiology Research Center (EPICARE) of Wake Forest University School of Medicine (Winston Salem, NC) using General Electric (GE) 12-SL software (GE, Milwaukee, WI) running under GE MUSE and Magellan Research Work Station.

The UKB digital ECGs were reported with five microvolts per least significant bit (uV/LSB). Median ECGs in CHS were reported with one uV/LSB, and in MESA, with 4.88. They were both rescaled to five uV/LSB prior to prediction from the ECG-AI model.

In CHS and MESA, the PTFV1 is defined as the duration (milliseconds) of the downward deflection (terminal portion) of the P-wave in lead V1 multiplied by the absolute value of its amplitude (μV).

### Echocardiography

Left atrial (LA) strain was measured using archived echocardiograms originally acquired in 1989–1990 and 1994–1995^52^, which were digitized from Super VHS tapes between 2016 and 2018. LA reservoir strain was assessed from apical 4-chamber views using speckle-tracking echocardiography (TOMTEC v4.5), with the LA endocardial border manually traced and strain curves generated from well-tracked segments^53^. Analyses were performed by experienced readers with ECG gating, and all strain values were reported as positive absolute percentages relative to the ventricular cycle as the reference point. LA reservoir strain was defined as the peak average LA strain and was calculated as the difference between peak strain values and the reference point.

### ECG-AI model

ECG-AI models to predict CMR measured LAVmin, LAVmax and LAEF were trained in UKB using a deep convolutional neural network (CNN). Since the model aims to predict structural aspects of the heart that are the same in each beat, to reduce dimensionality, we chose the median beat waveform as the input to our model. The UKB sample was split into 45% training, 5% validation and a 50% hold-out testing data sets. A large testing split was selected to enable the evaluation of each LA metric’s association with clinical outcomes. Input to the model was an 8×600 matrix representing the eight non-derived leads and 600-point time series waveform.

The CNN core architecture is an inception network that comprises four inception modules followed by three fully connected layers. An initial 1D convolutional layer is applied to extract low-level features from the raw ECG data, followed by batch normalization to stabilize the activations. Next, the model employs a series of four Inception-style blocks. In each block, the network processes the features in three parallel branches using convolutional layers with different kernel sizes scaled by factors of 1.0, 1.5, and 2.0 relative to the base filter length; each branch is followed by batch normalization. The outputs from these branches are then concatenated along the feature axis to integrate information captured at multiple temporal scales. Next, a Global Average Pooling layer reduces the concatenated feature maps to a single vector, which is then fed through a sequence of fully connected (dense) layers. The model used the Adam optimizer^54^, and patience for early stopping^55^ was set to 3 epochs. Twenty models for each were trained using a random search with tuning parameters for the number and initial length of filters in the inception block, the batch size, the dense layer size, and the learning rate. The top five models for each outcome were ensembled using the mean of the predictions.

### Statistical analysis

Left atrial measures were evaluated as continuous variables to predict event outcomes. Each measure was centered and scaled to have a mean of zero and a standard deviation of one. Associations with outcomes were assessed using Cox proportional hazards models.

AF analyses were adjusted for age, sex, race, height, body mass index (BMI), systolic blood pressure (SBP), diastolic blood pressure (DBP), current smoking, antihypertensive medication use, prevalent diabetes mellitus (DM), heart failure, and myocardial infarction (MI). Individuals with prior AF or a pacemaker were excluded.

Ischemic stroke analyses were adjusted for age, sex, race, BMI, SBP, current smoking, antihypertensive medication use, DM, prevalent heart failure and MI. Individuals with prior AF, ischemic stroke, or a pacemaker were excluded.

Heart failure analyses were adjusted for age, sex, race, BMI, SBP, current smoking, antihypertensive medication use, DM, total cholesterol, high-density lipoprotein cholesterol and left ventricular ejection fraction (LVEF). LVEF was modeled as a continuous value in UKB and MESA and categorized as qualitatively normal or abnormal in CHS. Individuals with prior AF, heart failure, or a pacemaker were excluded.

In CHS and UKB race was modeled as self-report White or other, in MESA analyses were adjusted for self-reported race. All covariates were assessed at the time of CMR, echocardiogram, or ECG measurement.

Sensitivity analyses for associations of LA measures and AF, ischemic stroke, and heart failure were performed as above adding natural log transformed N-terminal pro-B-type natriuretic peptide (NT-proBNP) as a covariate.

### Risk score comparison

To evaluate the ability of ECG-AI to predict 5-year risk for AF and the AF related outcomes of ischemic and cardioembolic stroke assessed risk scores, we evaluated four models: (1) age and sex only (base), (2) base plus all three ECG-AI left atrial measures (ECG-AI), (3) CHARGE-AF risk score alone (CHARGE-AF), and (4) CHARGE-AF + ECG-AI (combined). Each model was trained in the test partition of UKB to predict time to AF using Cox proportional hazards models. Participants in the testing set of UKB were limited to those >=65 to align with the external populations. The weights for each variable were then used to create scores in the external cohort studies. The models were evaluated in the joint MESA-CHS sample to predict incident events within 5 years. Ninety-five percent confidence intervals were created using 10,000 bootstrap samples. CHARGE-AF was estimated on group of studies that included CHS and therefore the CHARGE-AF estimates may perform better in this analysis than in a fully independent sample.

### Subclinical atrial fibrillation prediction

We used the ECG-AI model to predict LA measurements at MESA Exam 5 (2010-12). The highest quintile for LAVImin and LAVImax, and the lowest quintile for LAEF were used to define a ‘high-risk’ stratum for each measurement. The CHARGE-AF score was calculated on participants at Exam 5 and the highest quintile of scores was used to define the ‘high-risk’ strata and the top quintile of NT-proBNP defined ‘high-risk’ population for NT-proBNP. In MESA at Exam 6 (2016-18) 1528 participants were screened using ZioPatch monitor that detects and stores up to 14 days of cardiac rhythm (Zio Patch XT, iRhythm Technologies, Inc, San Francisco, CA)^15^. Monitor-detected AF included both atrial fibrillation and atrial flutter; atrial fibrillation was defined as an irregularly irregular rhythm with absent P waves lasting at least 30 seconds. Participants were excluded if AF was detected by ECG at Exam 5, they were missing data for the CHARGE-AF score or NT-proBNP measurements. Subclinical AF was detected by the monitor in 86 participants. The high-risk strata were used to predict subclinical AF as detected by cardiac monitoring. Area under the curve for receiver-operator curves were reported along with sensitivity and specificity.

### Gradient-weighted class activation mapping

Gradient-weighted Class Activation Mapping (Grad-CAM) was used to identify the regions of the ECG most influential in making atrial measure predictions. The last convolutional layer of the model with the lowest validation loss was used to explore the model gradients. The average gradients were calculated and visualized as a heatmap for the ECGs with the 50 highest volume predictions, or 50 smallest predictions for LAEF. For volumes, the maximum gradient multiplied by the convolutional outputs were plotted; for LAEF the minimal gradient multiplied by the convolutional outputs were used. The average amplitude for lead II of the 50 visualized ECGs were plotted for context. In addition, the amplitude for lead II of the 50 ECG predicting the smallest atrial volumes, or 50 largest ejection fractions, were compared to the Grad-CAM intensity locations.

### Genome-wide association studies

In the UKB, 488,377 participants were genotyped for over 800,000 variants using DNA extracted from blood samples collected at the baseline examination^56^. Genotypes were directly called using the UK Biobank Axiom (Affymetrix, Santa Clara, California) and the UK Applied Biosystems Array. Imputation was carried out using the Haplotype Reference Consortium and UK10K+1000 Genomes reference panels. Variants that did not pass quality control checks were excluded from further UKB analyses. We performed Genome-wide Association Studies (GWAS) of the five ECG-AI left atrial traits: LAVmin, LAVmax, LAVImax, LAVImin and LAEF. Individuals with a prior history of AF, a pacemaker or non-European ancestry were excluded. Analyses were performed using a linear mixed model to account for relatedness using the GENESIS package^57^. To improve statistical power and control for Type I errors with a non-normal phenotype distribution, we implemented a fully adjusted two-stage procedure for rank-normalization when fitting the null model^58^. The residuals from a regression model adjusting for age at MRI, sex, ten principal components of ancestry, and an indicator for UK BiLEVE or UK Biobank Axiom array were rank-normalized. Transformed residuals were then fit in a null model adjusting for the same fixed effects and a sparse genetic relationship matrix (GRM). The GRM was constructed from the KING kinship coefficients limited to third degree relatives from UKB. The resulting null model was used to perform genome-wide score tests of genetic association for all individual variants with minor allele frequency (MAF) > 0.005. We used genomic control factor lambda and the regression intercept from LD Score regression (LDSC) v1.0.1 ^59,60^ to assess inflation due to population stratification or polygenicity. We estimated the trait heritability (*h*^2^) using the LDSC regression slope. For each GWAS, index variants were identified for each locus by iteratively taking the variant with the lowest p-value and excluding other variants within a 500 kb window. Nearest genes were annotated using GENCODE v26^61^. We further annotated these index SNPs by identifying any AF index SNP from the 142 index SNPs from Neilson et. al^24^ within 500 kb.

### Genetic trait correlations

We used LDSC to estimate genetic correlations between 11 clinically related outcomes and risk factors and (1) the CMR atrial traits and (2) the ECG-AI predicted traits using default settings, which leave the intercept unconstrained. Trait GWAS were restricted to European ancestry individuals and limited to variants with MAF > 0.01.

Outcome trait GWAS used the largest available European ancestry meta-analyses for AF^24^, ischemic and cardioembolic stroke^62^, heart failure ^63^, Alzheimer’s disease^64^ and type II diabetes^65^. Correlation for related traits and risk factors used large publicly available GWAS of body mass index^66^, SBP and DBP^67^, PR interval,^68^ QT interval^69^. To evaluate LVM and LVEF, we performed GWAS of these traits using the measures derived from the Bai et al. segmentation method.^16^ LVM was indexed to body surface area. Analyses were restricted to individuals of European ancestry and variants with a minor allele frequency > 0.005. GWAS analyses used the methods described above, adjusting for age at imaging visit sex, 10 principal components of ancestry, UK BiLEVE or UK Biobank Axiom array, and imaging center.

## Notes

### Competing Interest Statement

Dr. Tison is a consultant to Viz.ai.

### Funding Statement

UK Biobank: This research has been conducted using the UK Biobank Resource under Application Number #40713. The authors would like to thank the participants and researchers involved in creating and maintaining the UK Biobank as a valuable resource for health-related research.
Multi-Ethnic Study of Atherosclerosis: This research was supported by contracts 75N92020D00001, HHSN268201500003I, N01-HC-95159, 75N92020D00005, N01-HC-95160, 75N92020D00002, N01-HC-95161, 75N92020D00003, N01-HC-95162, 75N92020D00006, N01-HC-95163, 75N92020D00004, N01-HC-95164, 75N92020D00007, N01-HC-95165, N01-HC-95166, N01-HC-95167, N01-HC-95168 and N01-HC-95169 from the National Heart, Lung, and Blood Institute, grants UL1-TR-000040 and UL1-TR-001420 from NCATS, and UL1-RR-025005 from NCRR and R01-HL-127659, R01-HL-107577 and R01 HL127659 from NHLBI. The authors thank the other investigators, the staff, and the participants of the MESA study for their valuable contributions. A full list of participating MESA investigators and institutions can be found at http://www.mesa-nhlbi.org.
Cardiovascular Health Study: This Cardiovascular Health Study research was supported by NHLBI contracts HHSN268201200036C, HHSN268200800007C, HHSN268201800001C, N01HC55222, N01HC85079, N01HC85080, N01HC85081, N01HC85082, N01HC85083, N01HC85086, N01HC85084, N01HC35129, R01AG15928, R01AG20098, 75N92021D00006; and NHLBI grants U01HL080295, R01HL087652, R01HL103612, R01HL105756, R01HL120393, U01HL130114, and R01HL172803 with additional contribution from the National Institute of Neurological Disorders and Stroke (NINDS). Additional support was provided through R01AG023629 from the National Institute on Aging (NIA). A full list of principal CHS investigators and institutions can be found at CHS-NHLBI.org. The content is solely the responsibility of the authors and does not necessarily represent the official views of the National Institutes of Health.
JSF was supported by NIH grants HL142599, HL149706, and HL172495.

### Author Declarations

For the Multi-Ethnic Study of Atherosclerosis and the Cardiovascular Health Study (CHS), written informed consent was obtained from all participants, and the institutional review board at each study site approved the study. The UK Biobank database received ethical approval from the North-West Haydock Research Ethics Committee.

