## Supplement for "Deep Learning Prediction of Left Atrial Structure and Function from 12-lead Electrocardiograms"

**Supplemental Figure 1: Consort Diagram of UKB Analyses**

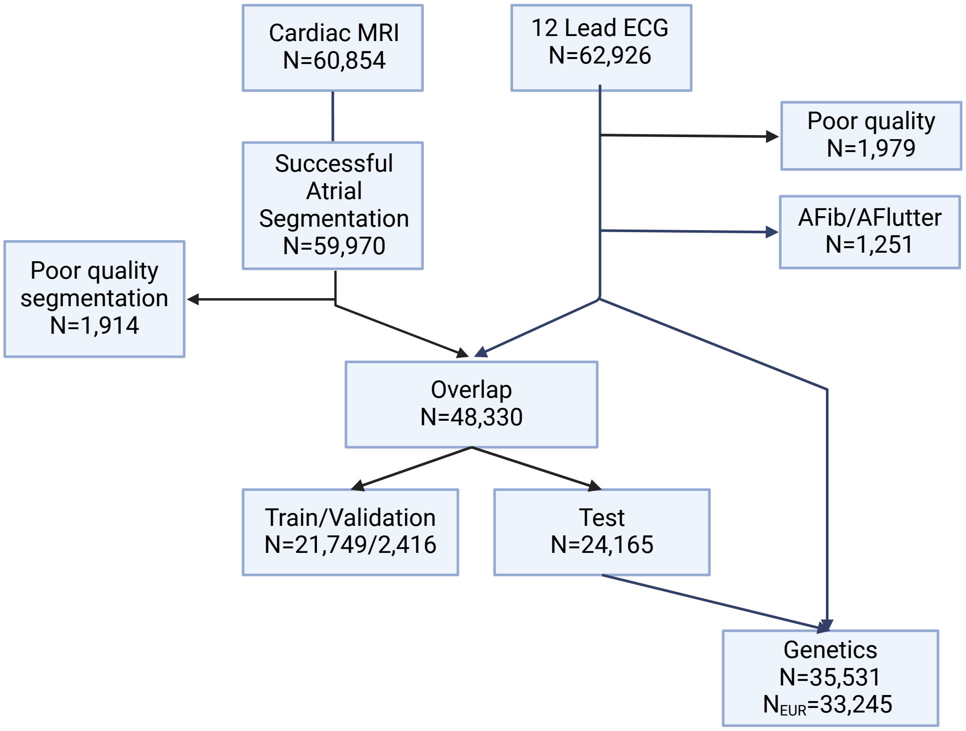


All cardiac MRIs were retrieved from UKB at the initial imaging visit and cardiac structure segmentation was attempted on all participants with imaging files for the short axis, long axis 2-chamber and 4-chamber view. Poor quality segmentation results were removed. All 12-lead ECGs from the same visit were downloaded and poor quality or ECGs with reported codes for atrial fibrillation or flutter were removed. The resulting 48,330 samples with both CMR and ECG were the ECG-AI model dataset. The UKB data were split into 45% training, 5% validation, and 50% testing datasets. Correlations with CMR volumes and UKB outcomes were performed in the Test dataset. Left atrial measures were predicted into the additional ECGs that did not have matching CMR data and were of European ancestry for genetics analyses.

**Supplemental Figure 2: Correlation Between Segmented CMR measures and ECG-AI Predicted Measures in MESA**

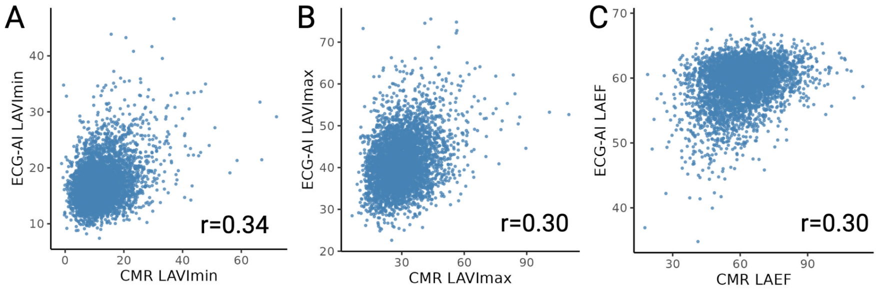

A
ECG-AI LAVImin
CMR LAVImin
r=0.34
B
ECG-AI LAVImax
CMR LAVImax
r=0.30
C
ECG-AI LAEF
CMR LAEF
r=0.30

**Supplemental Figure 3: Gradient-weighted Class Activation Mapping of ECG Features in Left Atrial Prediction Modeling**

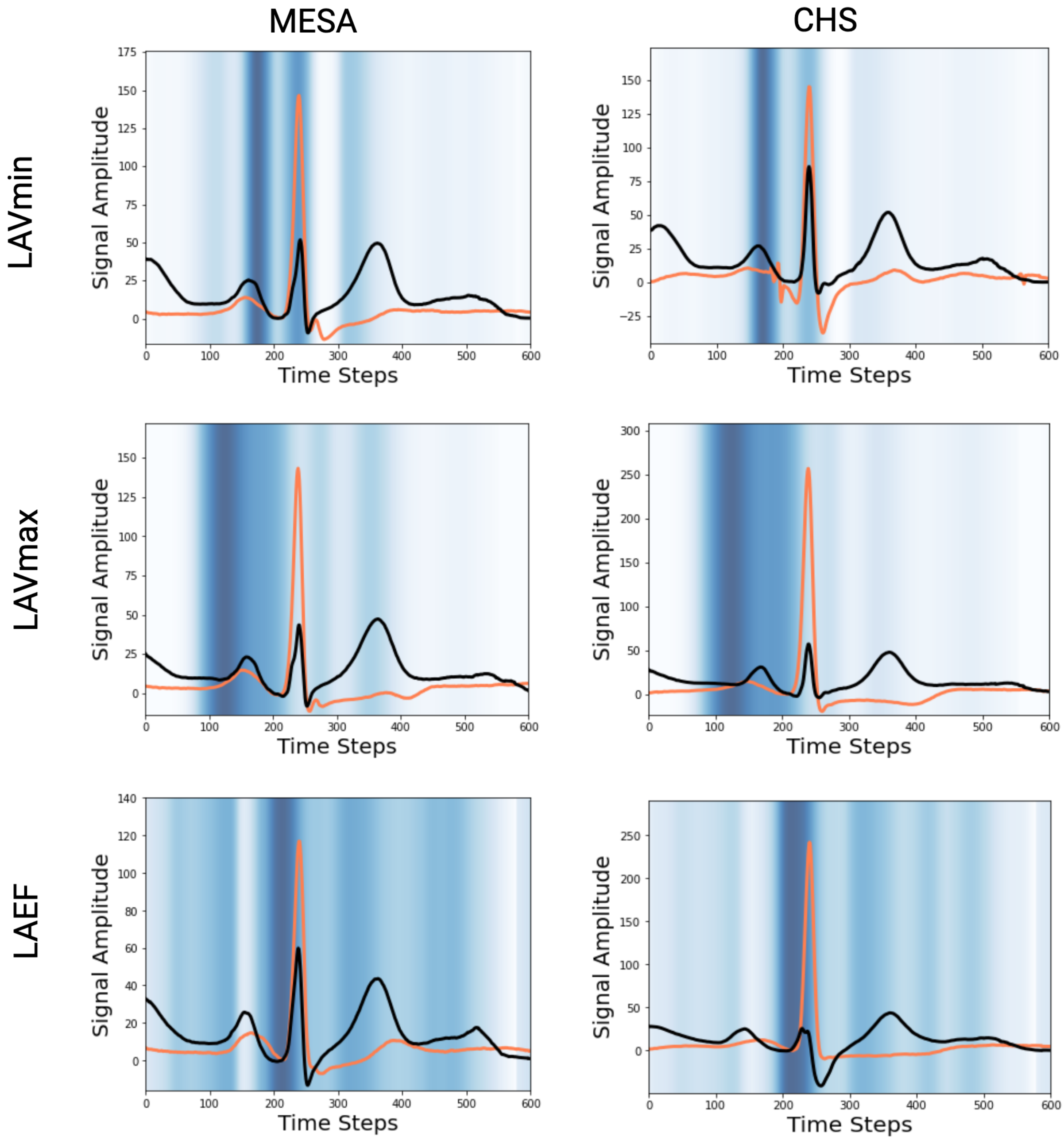


This figure shows the heatmaps for the electrocardiogram (ECG) region driving the predictions of the ECG-AI model across three cohorts. The blue intensity corresponds to feature importance, with the darkest blue signaling the highest importance. The 50 largest volumes (worse) for LAVmin and the 50 smallest ejection fractions for LAEF (worse) median beats are averaged and displayed in orange. For comparison, the average of the 50 smallest (volumes) or largest (LAEF) are plotted in black. For left atrial minimal volume (LAVmin), the model is primarily focused on the decline of the P-wave, while left atrial maximal volume (LAVmax) is focused on the proximal P-wave. In contrast, the left atrial ejection fraction (LAEF) signal is focused on the PQ segment, with consistent P-wave signaling intensities across the three cohort studies.

**Supplemental Figure 4: Manhattan Plot for ECG-AI predicted LA measures in UKB participants**

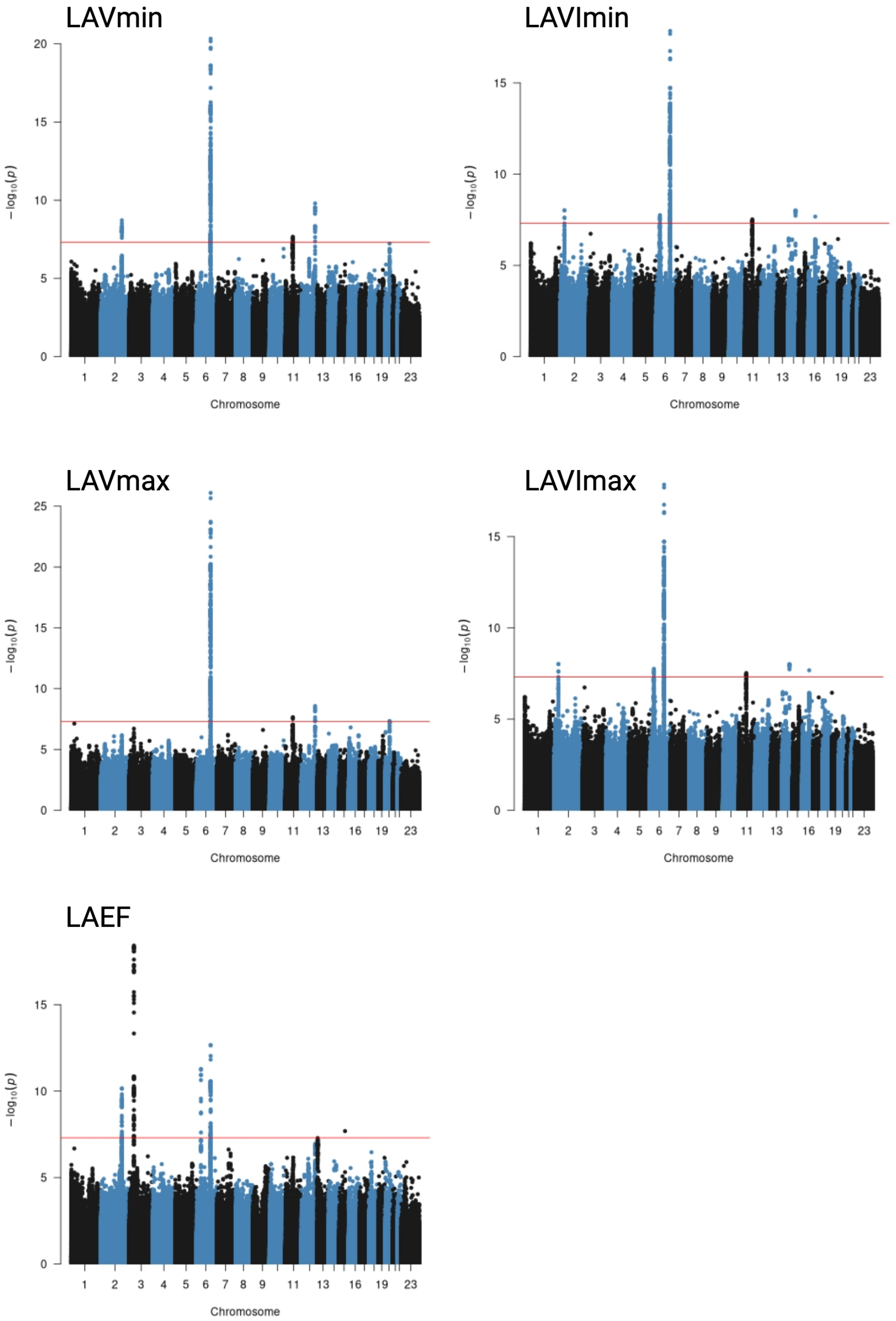

Each point represents a genetic variant. The x-axis represents the genomic chromosomal position, and the y-axis represents is the −log10(p value) of the genetic association. The red horizonal line represents the threshold for significance at 5e-8.
